# The Impact of Sampling Type, Frequency and Scale of Collection System on SARS-CoV-2 Quantification Fidelity

**DOI:** 10.1101/2021.07.07.21260158

**Authors:** Andrea D. George, Devrim Kaya, Blythe A. Layton, Kestrel Bailey, Christine Kelly, Kenneth J. Williamson, Tyler S. Radniecki

## Abstract

With the rapid onset of the COVID-19 pandemic, wastewater-based epidemiology (WBE) sampling methodologies for SARS-CoV-2 were often implemented quickly and may not have taken the unique drainage catchment characteristics into account. One question of debate is the relevance of grab versus composite samples when surveying for SARS-CoV-2 at various catchment scales. This study assessed the impact of grab versus composite sampling on the detection and quantification of SARS-CoV-2 in catchment basins with flow rates ranging from high-flow (wastewater treatment plant influent), to medium-flow (neighborhood-scale micro-sewershed), to low-flow (city block-scale micro-sewershed) and down to ultra-low flow (building scale). At the high-flow site, grab samples were reasonably comparable to 24-h composite samples with the same non-detect rate (0%) and SARS-CoV-2 concentrations that differed by 32% on the Log_10_ scale. However, as the flow rates decreased, the percentage of false-negative grab samples increased up to 44% and the SARS-CoV-2 concentrations of grab samples varied by up to 1-2 orders of magnitude compared to their respective composite sample concentrations. At the ultra-low-flow site, increased sampling frequencies down to every 5 min led to composite samples with higher fidelity to the SARS-CoV-2 load. Thus, composite sampling is superior to grab sampling, especially as flow decreases.

**Synopsis:** The need for composite sampling to generate reliable SARS-CoV-2 wastewater based epidemiology results increases as the collection basin scale decreases.

**Table of Content Art:** 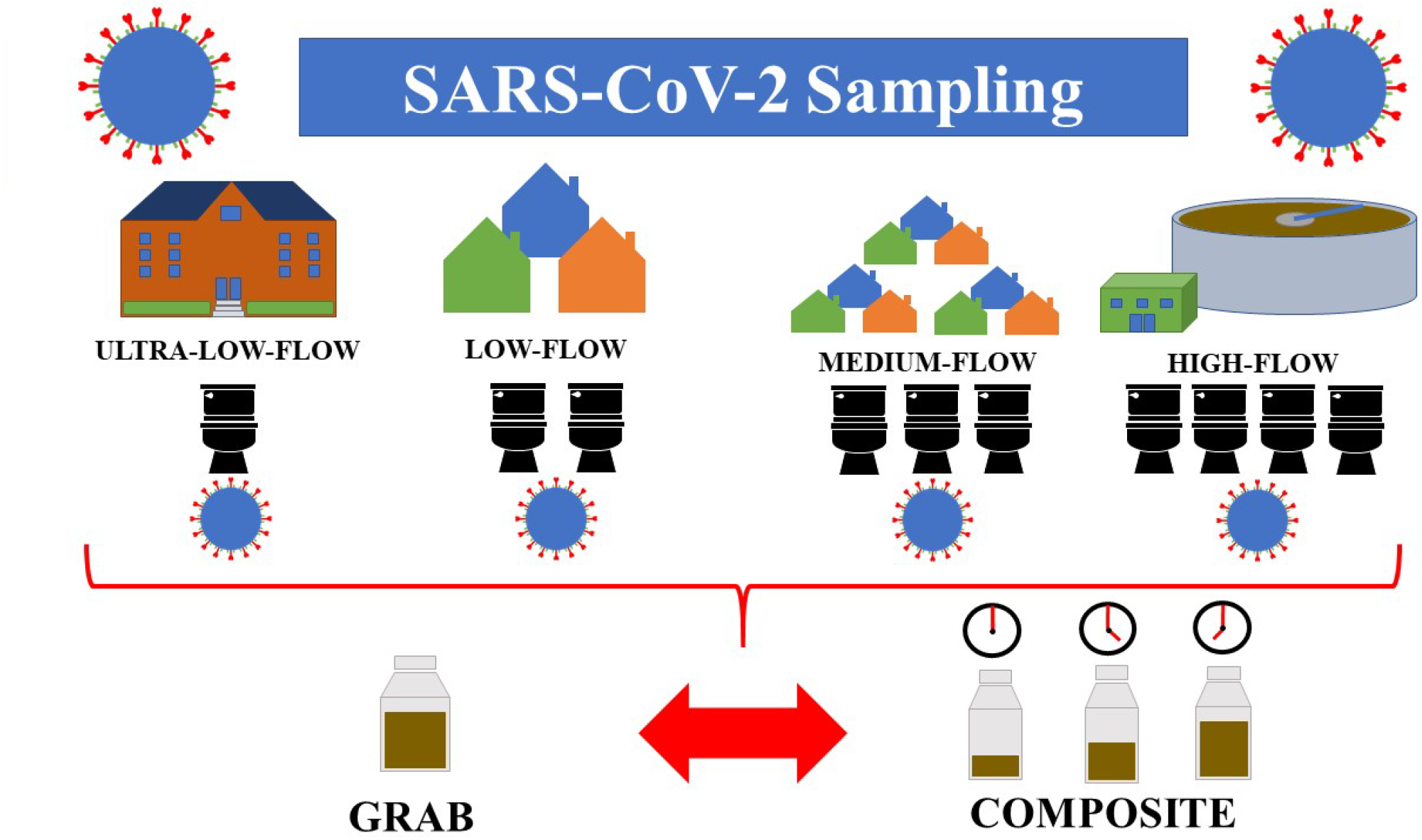

## Introduction

Since the start of the COVID-19 pandemic, wastewater-based epidemiology (WBE) has been widely used to track community SARS-CoV-2 viral burdens^1^. However, due to the temporal variability in SARS-CoV-2 loading into sewersheds, non-optimized sampling approaches in terms of type and/or frequency may introduce unintended biases. These include false negatives and gross over- or under-estimation of total daily viral loads.

Two of the most common WBE sampling approaches are grab and composite sampling. Composite sampling has been used to improve detection given the uncertainty of shedding rates and fluctuations in diurnal wastewater flowrate^1–5^. A downside to composite sampling is that it requires investing in costly and cumbersome equipment (*e*.*g*. autosamplers). Thus, grab sampling has also been used to monitor for SARS-CoV-2 in wastewater treatment plant (WWTP) influents and low-flow sewers leaving buildings^6^.

While cheaper, faster, and less laborious than composite sampling, there are concerns surrounding the accuracy of grab sampling due to the discrete and variable nature of SARS-CoV-2 inputs into the catchment. Likewise, the 1-h sampling frequency for creating 24-h composites is an industry standard that was developed for monitoring large catchment basins where there is significant time and flow for the dispersion of a target signal. However, it is unclear if this sampling frequency is adequate to accurately capture SARS-CoV-2 signals at much lower flow rates closer to the input source where time and flow for dispersion is minimal.

This study identifies the impact of sampling type and frequency on SARS-CoV-2 detection and quantification in wastewater samples collected from several catchment scales, ranging from influent at a WWTP to a cluster of buildings on a college campus. This study also provides insight into the temporal variability of SARS-CoV-2 concentrations under various flow regimes and how that may impact the interpretation of results generated by the grab and composite sampling approaches commonly used in WBE.

## Materials and Methods

### Sampling Process and Site Description

Three sites with low-, medium-, and high-flow rates in the Forest Grove, Oregon sewershed were selected for 24 h sampling. The low-flow site (0.42 m^3^/min average dry weather flow) serves 400 people in a small residential community while also receiving industrial discharge from three food processing plants with 24 h operations. The medium-flow site (2.65 m^3^/min average dry weather flow) receives most of its flow from a 2,200-person residential community with some small commercial businesses. The high-flow site (9.20 m^3^/min average dry weather flow) is the influent to the Forest Grove WWTP that serves approximately 40,000 residents and receives a mixture of industrial wastewater.

Hourly grab samples (200 mL) were taken from the low-, medium-, and high-flow sites with an ice-cooled 24-bottle ISCO 3700 autosampler (Teledyne ISCO, Lincoln, NE). Time-weighted composite samples were prepared by combining 10 mL of each hourly grab sample together. The SARS-CoV-2 concentrations for grab and time-weighted composite samples were quantified as described below.

The ultra-low-flow site served four college dormitory buildings, one of which was used to temporarily house COVID-19 infected students for convalescence. Grab samples (400 mL) were collected every 15 min for an 8 h period and every 5 min in the first 2 h. Time-weighted composites were calculated *in silico* by averaging the SARS-CoV-2 concentrations of the grab samples.

### Sample Concentration

Samples were stored at -20 °C for 1-50 d before concentration (**Table S1**). Frozen samples were thawed in a warm water bath (∼30°C) and concentrated by filtering samples (10-50 mL, depending on suspended solid content) through an electronegative mixed cellulose ester membrane filter (Whatman catalog no. 7141-104, Buckinghamshire, UK). Following filtration, filters were placed into 2-mL tubes containing 0.7-mm garnet beads and 1 mL DNA/RNA Shield (Zymo Research, Irvine, CA) and stored at -80 °C for 1-17 days (mean: 6 ± 4 days) until RNA extraction.

### RNA Extraction

All concentrated samples were thawed at room temperature (∼20 °C) and homogenized using a BioSpec Mini-Beadbeater 16 (BioSpec Products, Inc, Bartlesville, OK) for 2 min. Samples were kept on ice during RNA extraction. The beads and debris were centrifuged at 12,000 rcf for 1 min to remove debris. Lysate was transferred from each tube to a 96-well plate. RNA was extracted from 200 μL of lysate using the MagMAX Viral/Pathogen kit on a KingFisher automated instrument (ThermoFisher Scientific, Waltham, MA). Purified RNA was eluted in 30 µL in elution buffer provided with the extraction kit. Positive SARS-CoV-2 controls containing the E, N, ORF1ab, RdRP, and S genes and human RNAse P RNA (EDX SARS-CoV-2 Standard, Exact Diagnostics, Fort Worth, TX) and negative controls containing certified SARS-CoV-2-free human RNAse P RNA (EDX SARS-CoV-2 Negative, Exact Diagnostics, Fort Worth, TX) were included in each extraction plate. Extraction blanks of phosphate buffered saline (PBS) were included with every run as an extraction contamination control. Reverse transcriptase droplet digital PCR (RT-ddPCR) immediately followed RNA extraction and purification.

### Reverse Transcriptase Digital Droplet PCR

SARS-CoV-2 was quantified using a commercial triplex assay (2019-nCoV CDC ddPCR Triplex Probe Assay, Bio-Rad catalog no. 12008202) and the one-step RT-ddPCR Advanced Kit for Probes on the QX-200 ddPCR system (Bio-Rad, Hercules, CA). This assay uses the CDC’s N1 and N2 primers with RNAse P included as an internal control. The primer and probe sequences were published previously^7^. An automated droplet generator generated the droplets (mean: 12,534 (± 2114) droplets per reaction). Duplicate analyses were performed for each sample and control. No template controls were included on each plate. The one-step thermal cycling conditions were as follows: reverse transcription at 50 °C for 60 min; enzyme activation at 95 °C for 10 min; 40 cycles of denaturation at 94 °C for 30 s followed by annealing/extension at 55 °C for 60 s; enzyme inactivation at 98 °C for 10 min; and lastly a 4 °C hold for droplet stabilization, for a minimum of 30 min to a maximum of overnight. Finally, the amplification in the droplets was determined using the Bio-Rad droplet reader. All assay conditions were performed as specified in the Bio-Rad assay protocol^8^.

### Data Analysis

The QuantaSoft Analysis Pro software (Bio-Rad, Hercules, CA) was used to manually call droplet clusters for each target. R (version 4.0.2) with Rstudio Desktop (version 1.3.1056) and Microsoft Excel was used for all other analysis, and graphics were created with Microsoft Excel and ggplot2^9^. N1 and N2 concentrations showed good agreement and thus their geometric mean was used for all analyses (**Figure S1**).

### Quality Control and Error

In data analysis, samples were only accepted if the corresponding extraction blank, field blank, negative control, and no-template control (NTC) were all negative for SARS-CoV-2 assays (*i*.*e*., N1 and N2) and BCoV (in process recovery experiment determined below). Reactions with less than 6,000 droplets were treated as failures and repeated. If the targets (*i*.*e*., N1 or N2 for SARS-CoV-2, and BCoV) were amplified in at least three droplets per reaction, the reaction was accepted as positive. Limit of detection (LOD) was determined empirically to be 8 copies per reaction for N1 and 12 copies per reaction for N2. The SARS-CoV-2 concentration of the sample was calculated using Equation 1 in Supporting Information and the values for each sample (*i*.*e*., four values for SARS-CoV-2: two for each N1 and N2 targets, or two values for bovine coronavirus (BCoV) for duplicate samples) aggregated by a geometric mean. Non-detect values were replaced with half of the sample-specific limit of detection when calculating the mean. The sample-specific LOD was calculated from a conservative theoretical detection limit of 3 copies/reaction using Equation 1 in Supporting Information.

### Bovine coronavirus (BCoV) process recovery control

Similar to other studies, an attenuated vaccine strain of BCoV, was selected as a process recovery control due to its morphological and structural similarity to SARS-CoV-2^10,11^. BCoV solution was prepared from freeze-dried Calf Guard cattle vaccine (Bovine Rotavirus-Coronavirus Vaccine from Zoetis, NJ, USA) as described in the Supplemental Information^12^.

To determine process recovery efficiency, 5 µL of BCoV was added to 25 mL of wastewater (n=8) just prior to filtration. The BCoV recovery was calculated by dividing the quantity measured in wastewater samples to the quantity added to each wastewater sample prior to concentration. The mean BCoV recovery was 57 (± 4) %. Non-spiked wastewater samples (n=4) were also quantified for BCoV to assess background concentration. No BCoV was detected in non-spiked samples.

## Results and Discussion

### Effect of Sampling Type on Presence/Absence Detection of SARS-CoV-2

One objective of WBE is to use the presence or absence of SARS-CoV-2 signal in a wastewater sample to indicate the presence or absence of COVID-19 in the community^6,13^. To determine how the sampling approach affects the reliability of presence/absence determination at different drainage basin scales, grab and composite samples (collected over the same sampling period) were collected from 4 catchments ranging in size from an entire sewershed with high flow down to a micro-sewershed encompassing a few buildings with ultra-low flow. At all four flows, SARS-CoV-2 was detected in the composites at concentrations above LOD. Additionally, all grab samples at the high flow site (*i*.*e*. the WWTP) yielded SARS-CoV-2 concentrations greater than LOD, similar to a recent study comparing composites collected over 1-h and 24-h at a WWTP^2^. However, as wastewater flow decreased, the percent occurrence of grab sample non-detects increased from 0% at the high flowrate up to 43.5% and 40.6% at the low flow and ultra-low flow locations, respectively (**Table 1**).

**Table 1.**
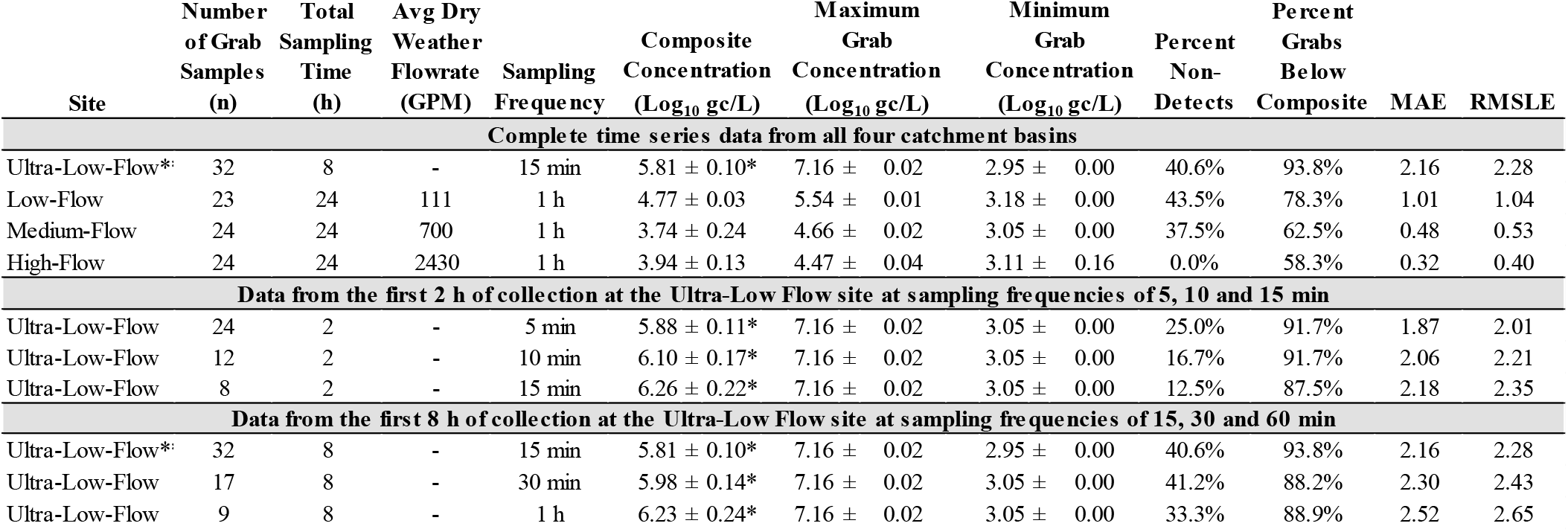
Summary of results at different flow rates and sampling frequencies. Ultra-Low-Flow composite concentrations (*) were calculated *in silico* from the corresponding grab samples. The Ultra-Low-Flow 15 min results (**) are shown twice for comparison purposes. MAE = mean absolute error. RMSLE = root mean squared log error.

| Site | Number of Grab Samples (n) | Total Sampling Time (h) | Avg Dry Weather Flowrate (GPM) | Sampling Frequency | Composite Concentration (Log<sub>10</sub> gc/L) | Maximum Grab Concentration (Log<sub>10</sub> gc/L) | Minimum Grab Concentration (Log<sub>10</sub> gc/L) | Percent Non-Detects | Percent Grabs Below Composite | MAE | RMSLE |
| --- | --- | --- | --- | --- | --- | --- | --- | --- | --- | --- | --- |
| Complete time series data from all four catchment basins |  |  |  |  |  |  |  |  |  |  |  |
| Ultra-Low-Flow* | 32 | 8 | - | 15 min | 5.81 ± 0.10* | 7.16 ± 0.02 | 2.95 ± 0.00 | 40.6% | 93.8% | 2.16 | 2.28 |
| Low-Flow | 23 | 24 | 111 | 1 h | 4.77 ± 0.03 | 5.54 ± 0.01 | 3.18 ± 0.00 | 43.5% | 78.3% | 1.01 | 1.04 |
| Medium-Flow | 24 | 24 | 700 | 1 h | 3.74 ± 0.24 | 4.66 ± 0.02 | 3.05 ± 0.00 | 37.5% | 62.5% | 0.48 | 0.53 |
| High-Flow | 24 | 24 | 2430 | 1 h | 3.94 ± 0.13 | 4.47 ± 0.04 | 3.11 ± 0.16 | 0.0% | 58.3% | 0.32 | 0.40 |
| Data from the first 2 h of collection at the Ultra-Low Flow site at sampling frequencies of 5, 10 and 15 min |  |  |  |  |  |  |  |  |  |  |  |
| Ultra-Low-Flow | 24 | 2 | - | 5 min | 5.88 ± 0.11* | 7.16 ± 0.02 | 3.05 ± 0.00 | 25.0% | 91.7% | 1.87 | 2.01 |
| Ultra-Low-Flow | 12 | 2 | - | 10 min | 6.10 ± 0.17* | 7.16 ± 0.02 | 3.05 ± 0.00 | 16.7% | 91.7% | 2.06 | 2.21 |
| Ultra-Low-Flow | 8 | 2 | - | 15 min | 6.26 ± 0.22* | 7.16 ± 0.02 | 3.05 ± 0.00 | 12.5% | 87.5% | 2.18 | 2.35 |
| Data from the first 8 h of collection at the Ultra-Low Flow site at sampling frequencies of 15, 30 and 60 min |  |  |  |  |  |  |  |  |  |  |  |
| Ultra-Low-Flow* | 32 | 8 | - | 15 min | 5.81 ± 0.10* | 7.16 ± 0.02 | 2.95 ± 0.00 | 40.6% | 93.8% | 2.16 | 2.28 |
| Ultra-Low-Flow | 17 | 8 | - | 30 min | 5.98 ± 0.14* | 7.16 ± 0.02 | 3.05 ± 0.00 | 41.2% | 88.2% | 2.30 | 2.43 |
| Ultra-Low-Flow | 9 | 8 | - | 1 h | 6.23 ± 0.24* | 7.16 ± 0.02 | 3.05 ± 0.00 | 33.3% | 88.9% | 2.52 | 2.65 |

Since all four sites had positive composites, the increase in grab sample non-detects at lower flow sites located closer to the input source suggests that the SARS-CoV-2 signal did not have enough time to spread out via dispersion mechanisms. This is reflected in the difference of the maximum and minimum SARS-CoV-2 concentrations observed at each location. This value was highest at the ultra-low flow site (4.21 Log_10_ gc/L) and decreased consistently as the flow increased, down to 1.36 Log_10_ gc/L at the high flow site (**Figure 1**). These results indicate grab samples may be acceptable at high-flow sites (*e*.*g*. WWTP influent) for presence/absence analyses, although this may change if the SARS-CoV-2 load is low in the community^2,14^. However, at low-flow or ultra-low flow sites (*e*.*g*. individual buildings), where SARS-CoV-2 signals appear in short bursts, composite sampling provides the most reliable information regarding signal presence.

**Figure 1.**
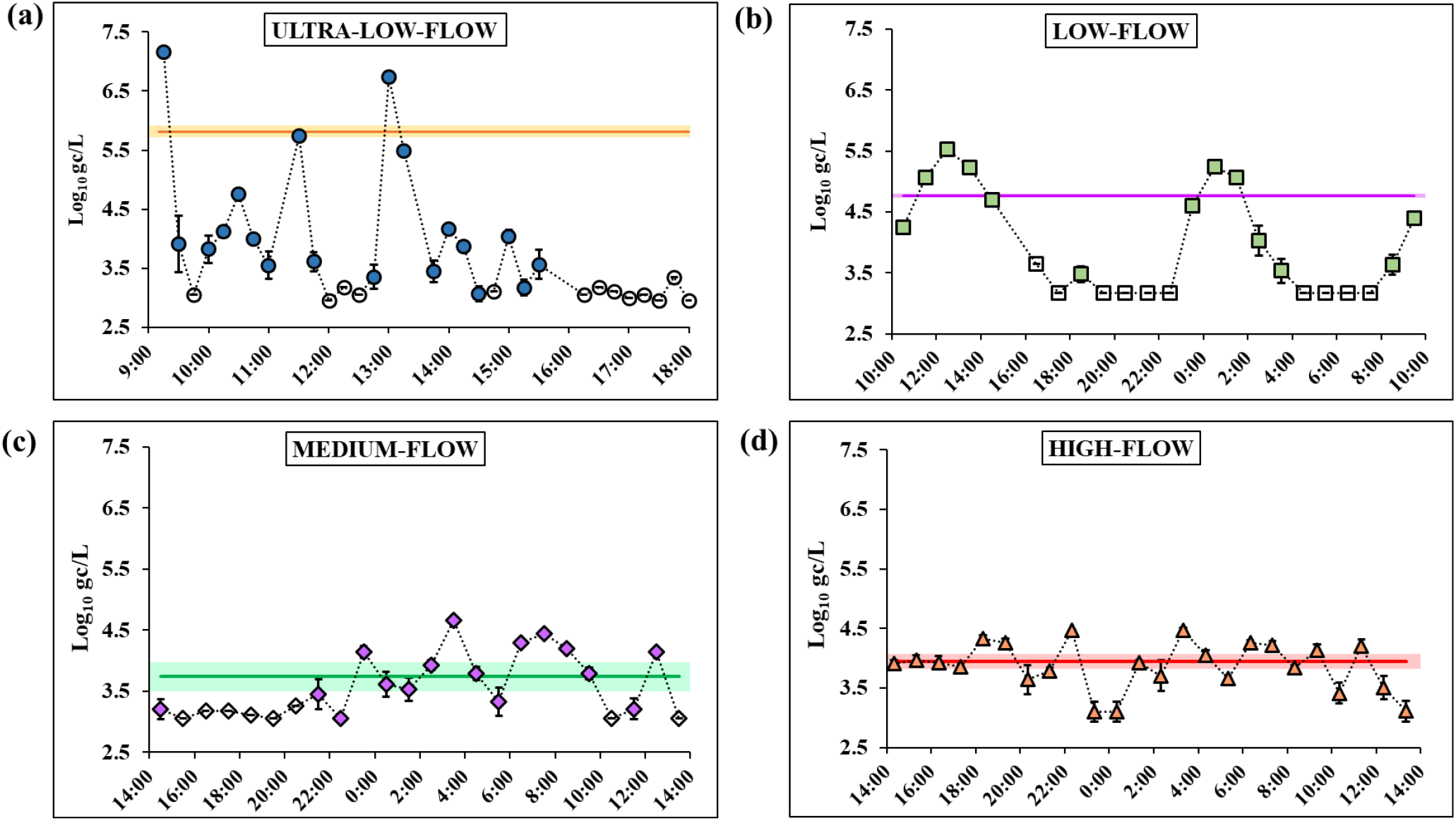
SARS-CoV-2 concentrations over time from grab samples collected from (a) ultra-low-flow (15-min sampling frequency), (b) low-flow, (c) medium-flow, and (d) high-flow sites. The solid line denotes the composite value for each time series. The error bars on the grab samples and the shaded range on the composite lines denote standard error. Non-detects are represented by open markers.

### Effect of Sampling Type on Quantification of SARS-CoV-2

Wastewater SARS-CoV-2 concentrations have been used to indicate community COVID-19 levels and to predict case rate changes^15,16^. Two different error metrics were employed to determine the impact of sampling approaches on SARS-CoV-2 quantification at different drainage basin scales. The first, mean absolute error (MAE), quantifies the average discrepancy between the SARS-CoV-2 concentrations observed in the grab samples versus the composite sample at each site (Equation 2, Supporting Information). The second, root mean square log error (RMSLE), applies more weight to outliers than the MAE, while also penalizing grab samples with concentrations below the composite value more than those above it (Equation 3, Supporting Information)^17,18^.

With both error metrics, the highest relative error between the grab samples and their respective composites was present at the ultra-low-flow site with MAE and RMSLE values of 2.16 and 2.28, respectively (**Table 1**). As the flow of the site increased, the difference between the grab and composite sample concentrations decreased, with the high-flow site having MAE and RMSLE values of 0.32 and 0.40, respectively. Thus, as the flow of the site increased, the relative error decreased and the grab samples became more representative of the composite value (**Figure S2**).

A recent study found that copies of SARS-CoV-2/100 mL of grab samples collected every 2 h from a WWTP influent were often within 50% of their respective 24-h flow-weighted composite copies/100 mL values^19^. Similar results were observed in the present study, with the high flow grab sample SARS-CoV-2 concentrations differing from their 24-h time-weighted composite concentration by an average of 32-40% of the log transformed value. Additionally, the grab sample SARS-CoV-2 concentrations at the high flow site were fairly well-distributed around the composite sample concentrations, with just over half (58.3%) of the grab sample values falling below the composite sample values.

However, this outcome was not exhibited at the other three sites with lower flow. Grab sample SARS-CoV-2 concentrations differed from their time-weighted composite sample values by around 1-2 orders of magnitude at the low-flow and ultra-low-flow site. Additionally, the distribution of the grab sample values around the composite sample concentrations was no longer approximately symmetrical (**Figure 1**). At the low-flow and ultra-low-flow sites, 78-94% of the grab sample concentrations were below the composite sample SARS-CoV-2 concentration (**Table 1**).

These results indicate that while grab samples may provide fairly representative SARS-CoV-2 concentrations at high flow sites (*e*.*g*. a WWTP influent), they fail to provide representative SARS-CoV-2 concentrations at lower flow sites (*e*.*g*. buildings) and may lead to over- or under-estimates of viral burden. This high level of variability at lower flow sites closer to the input source is indicative of limited SARS-CoV-2 dispersion by that point in the conveyance system, and highlights the need for composite sampling at such locations.

### Effect of Sampling Frequency on Quantification of SARS-CoV-2

The industry standard for creating 24-h time-weighted wastewater composites is to collect samples every hour, while recent studies have used a higher sampling frequency, ranging from 10 to 30 min, to monitor building-scale catchments^13,20^. Given the high variability of SARS-CoV-2 concentrations at the ultra-low-flow site (**Figure 1a**), questions arise regarding the optimal sampling frequency for building-scale catchments. To help answer these, the sampling frequency at the ultra-low-flow site was increased to every 5 min for the first 2 h (**Figure S3**) followed by a 15 min sampling frequency for the next 6 h of collection (**Figure 1a**).

For error metric analyses of sampling frequency over the first 2 h, the 5 min grab sample SARS-CoV-2 concentrations were used as the standard that all three sampling frequencies (5, 10, and 15 min) composite SARS-CoV-2 concentrations were compared against (**Figure S4**). This was done under the assumption that the 5 min grab samples are as close to continuous sampling as could be reasonably obtained and therefore would provide the most accurate reflection of the SARS-CoV-2 load during that time. Likewise, for error metric analyses for sampling frequency over the entire 8 h, the 15 min grab sample SARS-CoV-2 concentrations were used as the standard that the 15, 30, and 60 min sampling frequency composite SARS-CoV-2 concentrations were compared against (**Figure S5**).

For both time ranges (2 h and 8 h), the MAE and RMSLE increased with decreasing sampling frequency, with the highest values obtained from the composites from hourly sampling (**Table 1**). This demonstrates that reduced sampling frequency can result in the composite sample capturing less of the temporal variation of SARS-CoV-2 loads and reduces the accuracy of the composite sample’s SARS-CoV-2 concentration. Thus, at ultra-low-flow sites, it is advisable to increase the sampling frequencies to the greatest extent practical.

### Recommendations for Sampling Plan Design

Multiple factors should be considered in selecting the site-specific sampling method to optimize data collection. Among those factors, the scale of the catchment appears to strongly influence the sampling plan design by increasing the need for composite sampling at lower scales of flow. Additionally, the sampling frequency should also be considered, with special efforts to increase frequency at the individual building/campus level. Neither of these factors exist outside monetary and equipment restrictions; therefore, consideration should be given to the type of information desired to be obtained from the sampling (*e*.*g*., presence/absence versus quantitative viral concentrations) for better utilization and interpretation of the results.

## Data Availability

All data referred to in the manuscript in located within the manuscript.

## Abbreviations

BCoV: Bovine Corona Virus
LOD: Limit of Detection
MAE: Mean Average Error
NTC: No Template Control
rcf: Relative Centrifugal Field
RMSLE: Root Mean Squared Log Error
RT-ddPCR: Reverse Transcription Droplet Digital Polymerase Chain Reaction
WBE: Wastewater-based Epidemiology
WWTP: Wastewater Treatment Plant

## Acknowledgements

This work was supported by the National Science Foundation RAPID #2027679. Clean Water Services participated in this research with partial funding from the CARES Act through the Washington County Cities and Special District Assistance program. We wish to acknowledge the assistance with sampling from Scott Mansell, Jason Cook and Jacob DeMartino of Clean Water Services.

## Disclosures

The authors declare no competing financial interest.

## Supporting Information

### Equation S1. Conversion of copies/reaction to copies/L

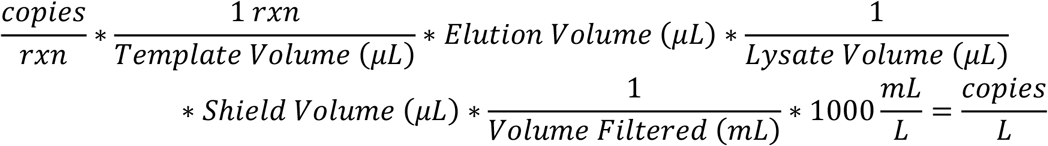

### Equation S2. Mean Absolute Error (MAE)

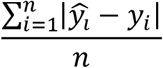

Where *ŷ*_*i*_ = observed value (log-transformed)

*y*_*i*_ = expected value (log-transformed)

n = number of observations

Please note that the interpretation of the MAE requires the recognition of the use of log-transformed values. The log-transformation of the concentration establishes the metric as one of relative rather than absolute error (as the name may suggest), since log rules dictate that there is a division of the grab by the composite within the MAE calculation. Thus, the resulting MAE is a percent error not an absolute error.

### Equation S3. Root Mean Square Log Error (RMSLE)

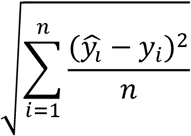

Where *ŷ*_*i*_ = observed value (log-transformed)

*y*_*i*_ = expected value (log-transformed)

*n* = number of observations

Please note that the interpretation of the RMSLE requires the recognition of the use of log-transformed values. The log-transformation of the concentration establishes the metric as one of relative rather than absolute error (as the name may suggest), since log rules dictate that there is a division of the grab by the composite within the RMSLE calculation. Thus, the resulting RMSLE is a percent error not an absolute error.

#### Bovine coronavirus (BCoV) stock solution preparation

BCoV solution was prepared from freeze-dried Calf Guard cattle vaccine (Bovine Rotavirus-Coronavirus Vaccine from Zoetis, NJ, USA) after rehydrating in 3 mL of sterile diluent provided with the vaccine. Aliquots (100 μL) of stock solution were stored at -20 ºC. Each aliquot was used for a maximum of two freeze-thaw cycles. To determine the stock concentration, 10 µL BCoV stock was added to 390 µL PBS. From this mixture, 200 μL were extracted as described above. The extracted BCoV RNA was serially diluted (1:10) in nuclease-free water for six dilutions and run in duplicate using a previously published BCoV assay by following the one-step RT-ddPCR procedure as described in the text^12^. Stock concentration of BCoV was around 2.3 x 106 gc/μL.

**Figure S1.**
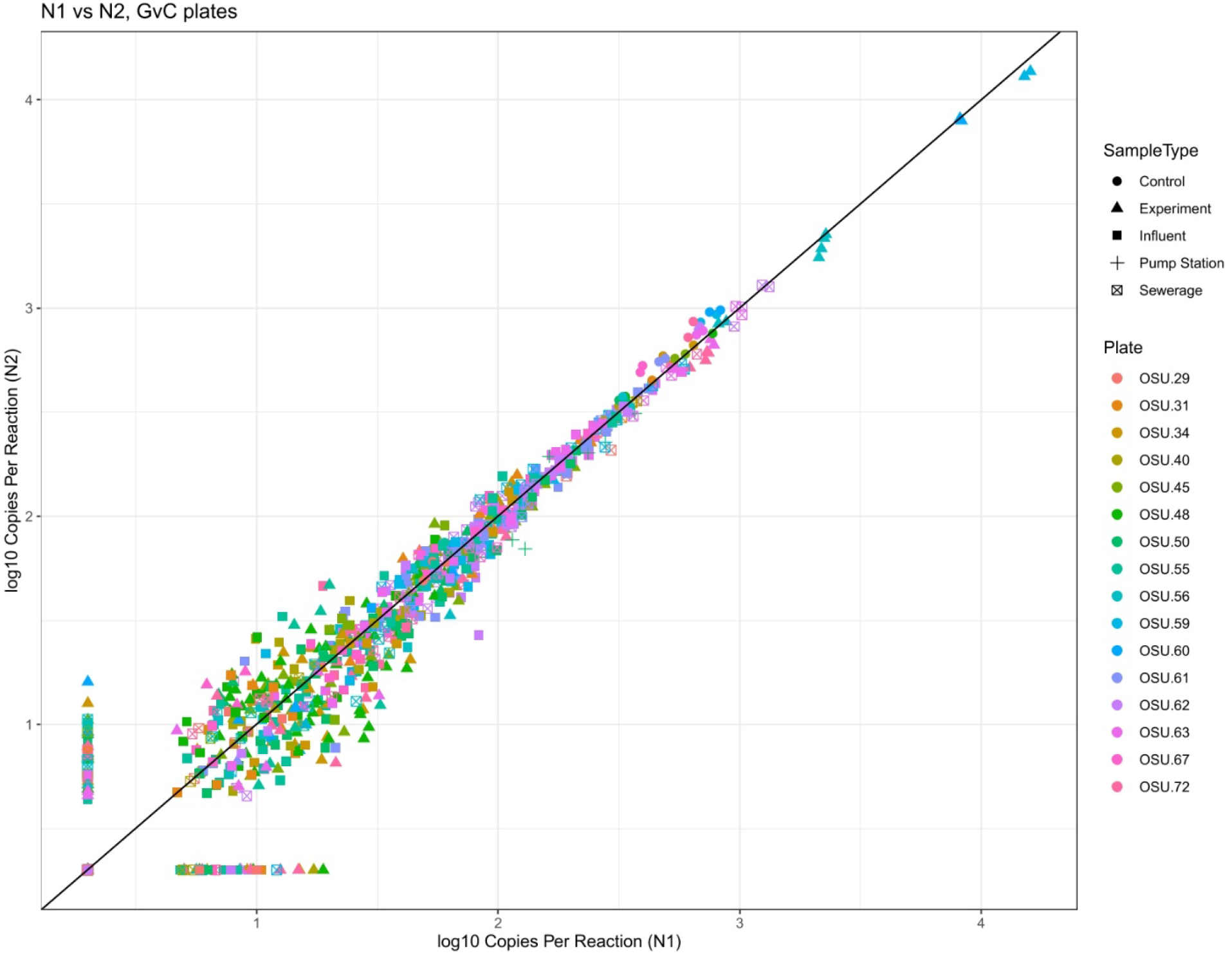
The N1 target concentrations (log_10_ copies per reaction) plotted against the N2 target concentration (log_10_ copies per reaction) in each reaction (n=1,328) in every plate containing samples utilized in this study. Non-detects were replaced with 3 copies per reaction.

**Figure S2.**
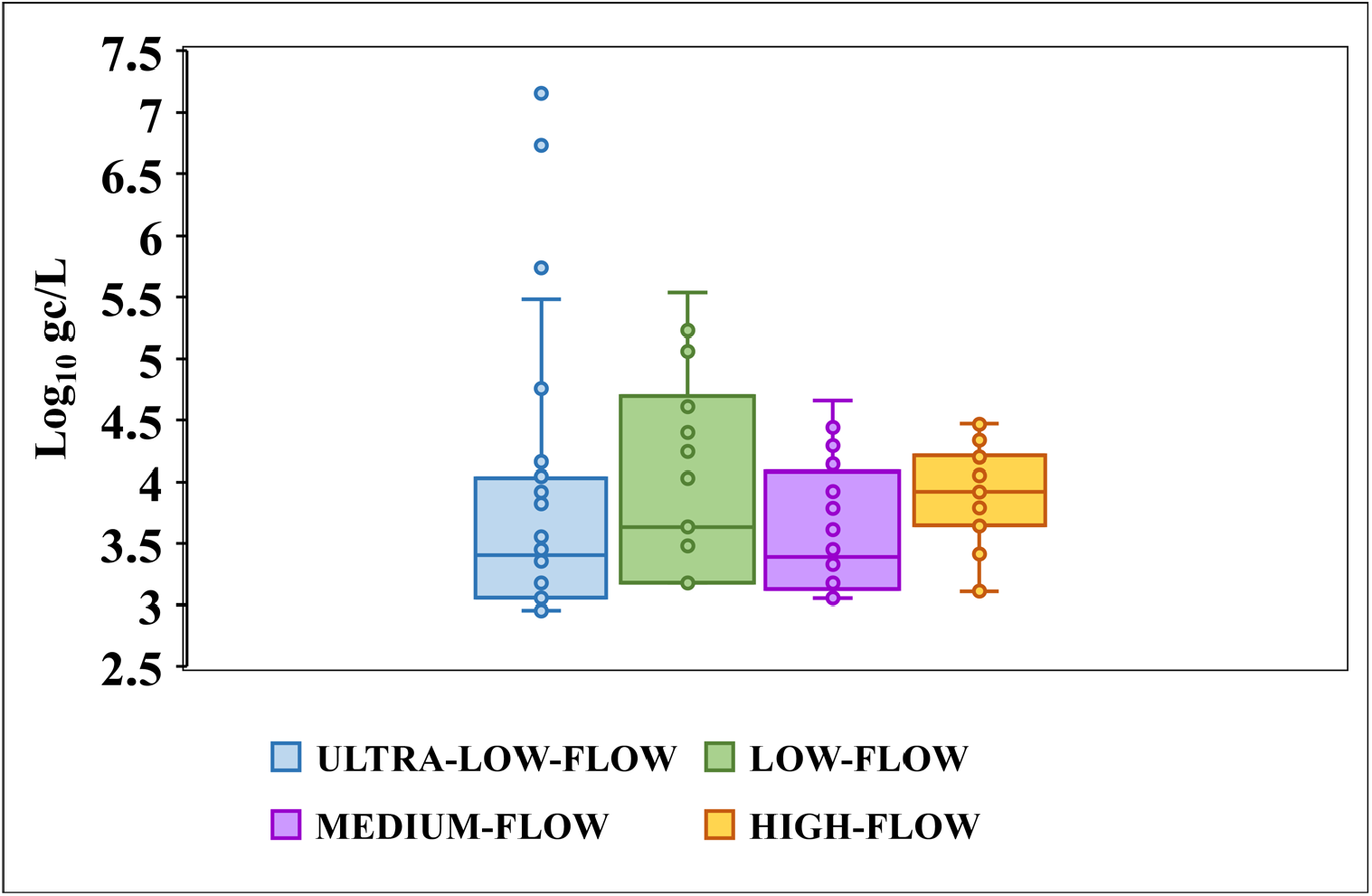
The boxplot exhibiting the distribution of viral concentrations (log_10_ gene copies/liter) of the grab samples at ultra-low-, low-, medium-, and high-flow sites. The size of the box has a negative correlation with flowrate. The interquartile range (IQR) decreases with increasing flow except at the ultra-low-flow site with the 15-minute sampling frequency (ultra-low-flow = 0.9, low-flow=1.5 log units, medium-flow=0.8 and the high-flow=0.5).

**Figure S3.**
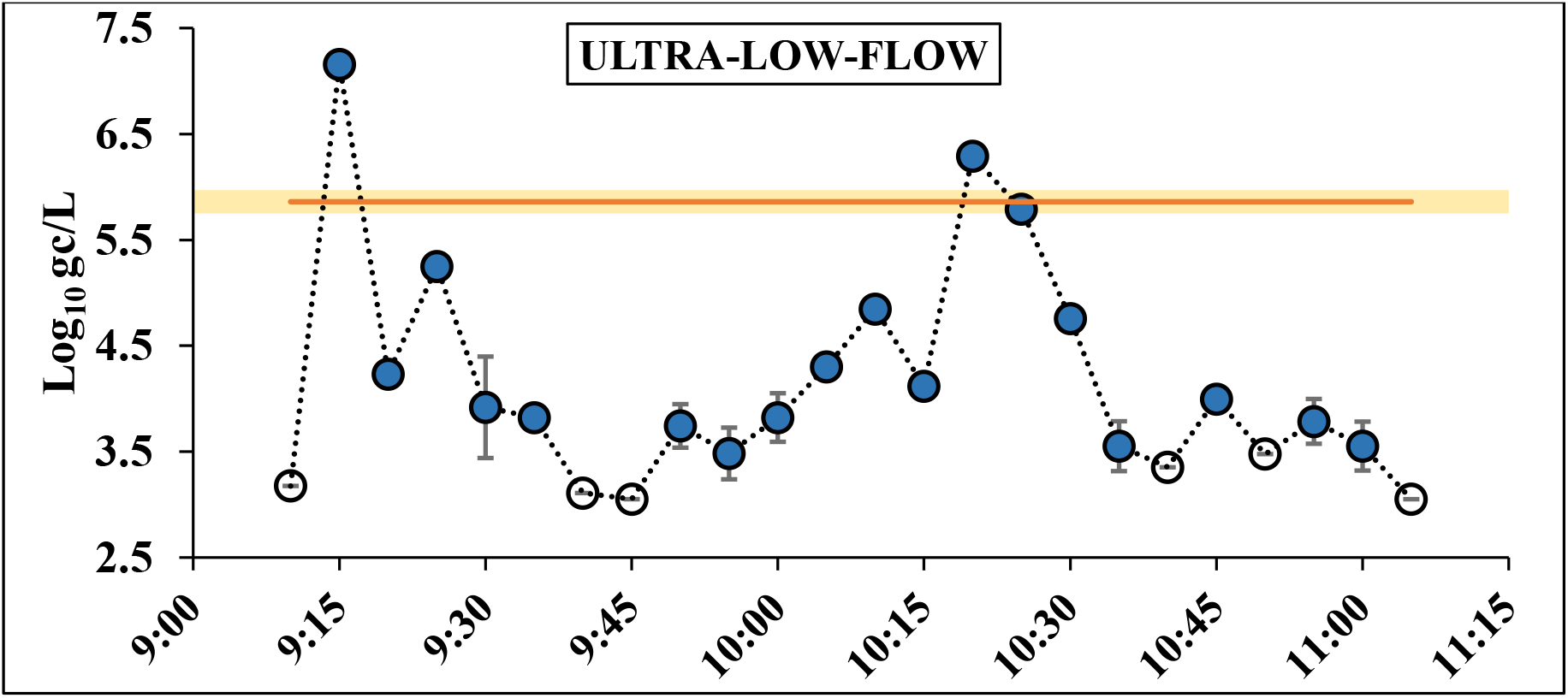
Variation in SARS-CoV-2 concentrations (log_10_ gene copies per liter, gc/L) over time in grab samples collected from the ultra-low-flow site over 5 min intervals in the first 2 h. The composite sample for the campus site was created digitally using the respective grab samples collected at 5 min intervals. The error bars on the grab samples and the shaded range on the composite lines denote standard error. Non-detects are represented by open markers.

**Figure S4.**
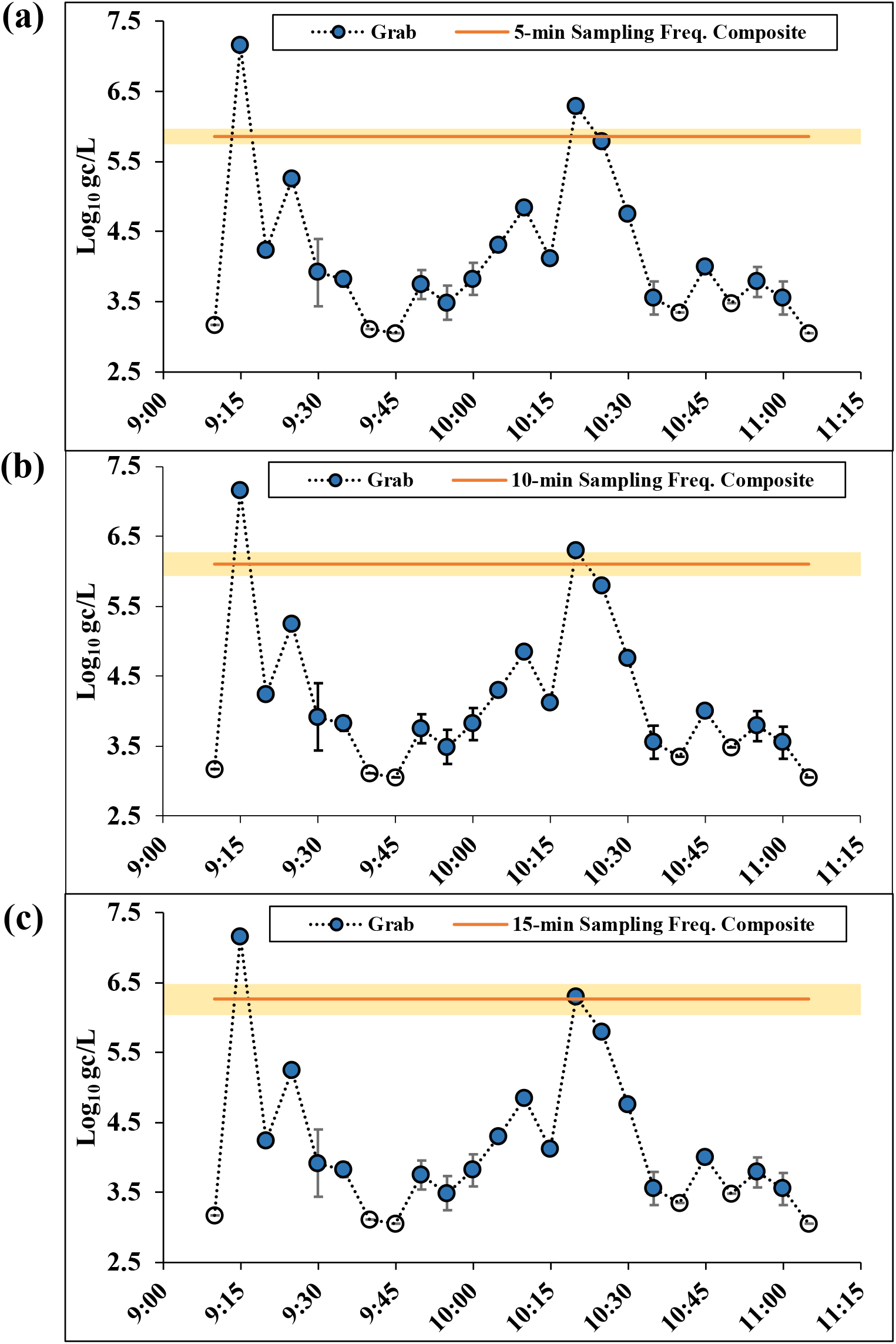
The 5-min grab samples collected in the first 2 h of sample collection plotted with a solid line indicating the (a) 5-min sampling frequency composite, (b) 10-min sampling frequency composite, and (c) 15-min sampling frequency composite. The error bars on the grab samples and the shaded range on the composite lines denote standard error. Non-detects are represented by open markers.

**Figure S5.**
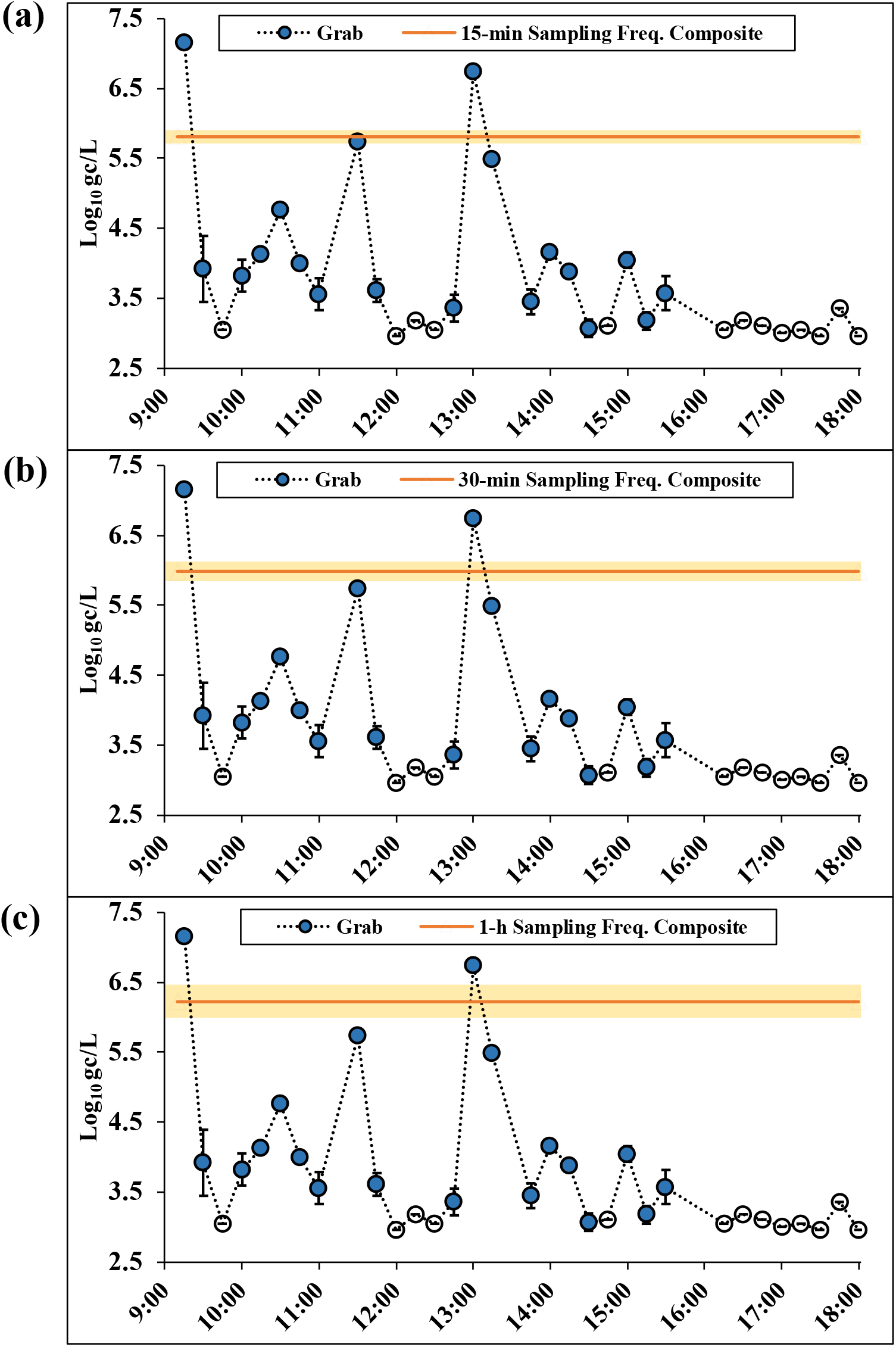
The 15-min grab samples collected in 8 h of sample collection plotted with a solid line indicating the (a) 15-min sampling frequency composite, (b) 30-min sampling frequency composite, and (c) 1-h sampling frequency composite. The error bars on the grab samples and the shaded range on the composite lines denote standard error. Non-detects are represented by open markers.

**Table S1.** The storage time, temperature, and volume concentrated for each of the raw wastewater samples collected from the four sites.

| LOCATION | HOUR | TEMP<br>STORED<br>(°C) | DAYS<br>STORED | VOLUME<br>FILTERED<br>(mL) |
| --- | --- | --- | --- | --- |
| Ultra-Low Flow | 9:10 | -20 | 50 | 30 |
| Ultra-Low Flow | 9:15 | -20 | 9 | 30 |
| Ultra-Low Flow | 9:20 | -20 | 50 | 35 |
| Ultra-Low Flow | 9:25 | -20 | 50 | 10 |
| Ultra-Low Flow | 9:30 | -20 | 9 | 30 |
| Ultra-Low Flow | 9:35 | -20 | 50 | 50 |
| Ultra-Low Flow | 9:40 | -20 | 50 | 35 |
| Ultra-Low Flow | 9:45 | -20 | 9 | 40 |
| Ultra-Low Flow | 9:50 | -20 | 50 | 30 |
| Ultra-Low Flow | 9:55 | -20 | 50 | 35 |
| Ultra-Low Flow | 10:00 | -20 | 9 | 30 |
| Ultra-Low Flow | 10:05 | -20 | 50 | 30 |
| Ultra-Low Flow | 10:10 | -20 | 50 | 40 |
| Ultra-Low Flow | 10:15 | -20 | 9 | 30 |
| Ultra-Low Flow | 10:20 | -20 | 50 | 10 |
| Ultra-Low Flow | 10:25 | -20 | 50 | 30 |
| Ultra-Low Flow | 10:30 | -20 | 9 | 30 |
| Ultra-Low Flow | 10:35 | -20 | 50 | 30 |
| Ultra-Low Flow | 10:40 | -20 | 50 | 20 |
| Ultra-Low Flow | 10:45 | -20 | 9 | 35 |
| Ultra-Low Flow | 10:50 | -20 | 50 | 15 |
| Ultra-Low Flow | 10:55 | -20 | 50 | 30 |
| Ultra-Low Flow | 11:00 | -20 | 9 | 30 |
| Ultra-Low Flow | 11:05 | -20 | 50 | 40 |
| Ultra-Low Flow | 11:30 | -20 | 24 | 40 |
| Ultra-Low Flow | 11:45 | -20 | 24 | 30 |
| Ultra-Low Flow | 12:00 | -20 | 24 | 50 |
| Ultra-Low Flow | 12:15 | -20 | 11 | 30 |
| Ultra-Low Flow | 12:30 | -20 | 11 | 30 |
| Ultra-Low Flow | 12:45 | -20 | 11 | 30 |
| Ultra-Low Flow | 13:00 | -20 | 11 | 45 |
| Ultra-Low Flow | 13:15 | -20 | 24 | 30 |
| Ultra-Low Flow | 13:45 | -20 | 24 | 50 |
| Ultra-Low Flow | 14:00 | -20 | 24 | 40 |
| Ultra-Low Flow | 14:15 | -20 | 24 | 35 |
| Ultra-Low Flow | 14:30 | -20 | 24 | 50 |
| Ultra-Low Flow | 14:45 | -20 | 31 | 35 |
| Ultra-Low Flow | 15:00 | -20 | 31 | 30 |
| Ultra-Low Flow | 15:15 | -20 | 31 | 40 |
| Ultra-Low Flow | 15:30 | -20 | 31 | 30 |
| Ultra-Low Flow | 16:15 | -20 | 31 | 40 |
| Ultra-Low Flow | 16:30 | -20 | 31 | 30 |
| Ultra-Low Flow | 16:45 | -20 | 31 | 35 |
| Ultra-Low Flow | 17:00 | -20 | 31 | 45 |
| Ultra-Low Flow | 17:15 | -20 | 31 | 40 |
| Ultra-Low Flow | 17:30 | -20 | 31 | 50 |
| Ultra-Low Flow | 17:45 | -20 | 31 | 20 |
| Ultra-Low Flow | 18:00 | -20 | 31 | 50 |
| Low-Flow | 1 | -20 | 12 | 40 |
| Low-Flow | 2 | -20 | 12 | 40 |
| Low-Flow | 3 | -20 | 12 | 30 |
| Low-Flow | 4 | -20 | 12 | 30 |
| Low-Flow | 5 | -20 | 12 | 35 |
| Low-Flow | 6 | -20 | 12 | 40 |
| Low-Flow | 7 | -20 | 12 | 25 |
| Low-Flow | 8 | -20 | 12 | 40 |
| Low-Flow | 9 | -20 | 12 | 40 |
| Low-Flow | 10 | -20 | 12 | 40 |
| Low-Flow | 11 | -20 | 12 | 40 |
| Low-Flow | 12 | -20 | 12 | 45 |
| Low-Flow | 13 | -20 | 12 | 50 |
| Low-Flow | 14 | -20 | 12 | 50 |
| Low-Flow | 15 | -20 | 12 | 50 |
| Low-Flow | 16 | -20 | 12 | 50 |
| Low-Flow | 17 | -20 | 12 | 40 |
| Low-Flow | 18 | -20 | 12 | 50 |
| Low-Flow | 19 | -20 | 12 | 40 |
| Low-Flow | 20 | -20 | 12 | 40 |
| Low-Flow | 21 | -20 | 12 | 40 |
| Low-Flow | 22 | -20 | 12 | 40 |
| Low-Flow | 23 | -20 | 12 | 30 |
| Low-Flow | 24 | -20 | 12 | 40 |
| Low-Flow | Composite | -20 | 12 | 40 |
| Med-Flow | 1 | -20 | 15 | 20 |
| Med-Flow | 2 | -20 | 15 | 20 |
| Med-Flow | 3 | -20 | 15 | 20 |
| Med-Flow | 4 | -20 | 15 | 20 |
| Med-Flow | 5 | -20 | 15 | 20 |
| Med-Flow | 7 | -20 | 15 | 10 |
| Med-Flow | 8 | -20 | 15 | 30 |
| Med-Flow | 9 | -20 | 15 | 20 |
| Med-Flow | 10 | -20 | 15 | 30 |
| Med-Flow | 11 | -20 | 15 | 30 |
| Med-Flow | 12 | -20 | 15 | 30 |
| Med-Flow | 13 | -20 | 15 | 30 |
| Med-Flow | 14 | -20 | 15 | 20 |
| Med-Flow | 15 | -20 | 15 | 20 |
| Med-Flow | 16 | -20 | 15 | 30 |
| Med-Flow | 17 | -20 | 15 | 20 |
| Med-Flow | 18 | -20 | 15 | 20 |
| Med-Flow | 19 | -20 | 15 | 30 |
| Med-Flow | 20 | -20 | 15 | 30 |
| Med-Flow | 21 | -20 | 15 | 30 |
| Med-Flow | 22 | -20 | 15 | 30 |
| Med-Flow | 23 | -20 | 15 | 20 |
| Med-Flow | 24 | -20 | 15 | 20 |
| Med-Flow | Composite | -20 | 15 | 30 |
| High-Flow | 1 | -20 | 8 | 50 |
| High-Flow | 2 | -20 | 8 | 50 |
| High-Flow | 3 | -20 | 8 | 50 |
| High-Flow | 4 | -20 | 8 | 50 |
| High-Flow | 5 | -20 | 8 | 30 |
| High-Flow | 6 | -20 | 8 | 50 |
| High-Flow | 7 | -20 | 8 | 50 |
| High-Flow | 8 | -20 | 8 | 50 |
| High-Flow | 9 | -20 | 8 | 50 |
| High-Flow | 10 | -20 | 8 | 50 |
| High-Flow | 11 | -20 | 8 | 50 |
| High-Flow | 12 | -20 | 8 | 50 |
| High-Flow | 13 | -20 | 8 | 50 |
| High-Flow | 14 | -20 | 8 | 50 |
| High-Flow | 15 | -20 | 8 | 50 |
| High-Flow | 16 | -20 | 8 | 50 |
| High-Flow | 17 | -20 | 8 | 50 |
| High-Flow | 18 | -20 | 8 | 50 |
| High-Flow | 19 | -20 | 8 | 50 |
| High-Flow | 20 | -20 | 8 | 50 |
| High-Flow | 21 | -20 | 8 | 50 |
| High-Flow | 22 | -20 | 8 | 50 |
| High-Flow | 23 | -20 | 8 | 50 |
| High-Flow | 24 | -20 | 8 | 50 |
| High-Flow | Composite | -20 | 8 | 50 |

## Notes

### Competing Interest Statement

The authors have declared no competing interest.

### Author Declarations

Oregon State University's IRB has given this work an exemption.

